# Associations Between Physical Fitness and Brain Structure in Young Adulthood

**DOI:** 10.1101/2020.09.18.20196014

**Authors:** John R. Best, Elizabeth Dao, Ryan Churchill, Theodore D. Cosco

## Abstract

A comprehensive analysis of associations between physical fitness and brain structure in young adulthood is lacking, and further, it is unclear the degree to which associations between physical fitness and brain health can be attributed to a common genetic pathway or to environmental factors that jointly influences physical fitness and brain health. This study examined genotype-confirmed monozygotic and dizygotic twins, along with non-twin full-siblings to estimate the contribution of genetic and environmental factors to variation within, and covariation between, physical fitness and brain structure. Participants were 1065 young adults between the ages of 22 and 36 from open-access Young Adult Human Connectome Project (YA-HCP). Physical fitness was assessed by submaximal endurance (two-minute walk test), grip strength, and body mass index. Brain structure was assessed using magnetic resonance imaging on a Siemens 3T customized ‘Connectome Skyra’ at Washington University in St. Louis, using a 32-channel Siemens head coil. Acquired T1-weighted images provided measures of cortical surface area and thickness, and sub-cortical volume following processing by the YA-HCP structural FreeSurfer pipeline. Diffusion weighted imaging was acquired to assess white matter tract integrity, as measured by fractional anisotropy, following processing by the YA-HCP diffusion pipeline and tensor fit. Following correction for multiple testing, body mass index was negatively associated with fractional anisotropy in various white matter regions of interest (all |*z*| statistics > 3.9) and positively associated with cortical thickness within the right superior parietal lobe (z statistic = 4.6). Performance-based measures of fitness (i.e., endurance and grip strength) were not associated with any structural neuroimaging markers. Behavioral genetic analysis suggested that heritability of white matter integrity varied by region, but consistently explained >50% of the phenotypic variation. Heritability of right superior parietal thickness was large (∼75% variation). Heritability of body mass index was also fairly large (∼60% variation). Generally, 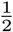 to 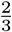 of the correlation between brain structure and body mass index could be attributed to heritability effects. Overall, this study suggests that greater body mass index is associated with lower white matter integrity, which may be due to common genetic effects that impact body composition and white matter integrity.

## Introduction

Associations between performance-based physical fitness and neuroimaging markers of brain health have been studied fairly extensively in the context of aging, and to a somewhat lesser extent in the context of childhood development. Previous studies have shown that physical fitness—most commonly cardiorespiratory fitness— positively correlates with gray matter thickness or volume in various regions of the cortex in children and older adults (Gordon et al., 2008;Chaddock-Heyman et al., 2015;Wood et al., 2016;Williams et al., 2017;Esteban-Cornejo et al., 2019;Cadenas-Sanchez et al., 2020) and with hippocampal volume in older adults (Erickson et al., 2009;Cole et al., 2020). Other research has shown associations between physical fitness—again typically cardiorespiratory fitness—and higher diffusion-based white matter integrity (Marks et al., 2011;Tseng et al., 2013;Oberlin et al., 2015;Ding et al., 2018) or reduced lesion volume in older adults (Sexton et al., 2016). In children, one study found a weak association between upper-body strength and white matter integrity in the frontal lobe (Rodriguez-Ayllon et al., 2020).

Relatedly, other research has looked at associations between body composition—most commonly body mass index (BMI)—and neuroimaging markers of brain health in youth and older adults. Higher BMI has been associated with smaller gray matter volume in various cortical and subcortical regions in adolescence (Kennedy et al., 2016) and late adulthood (Ho et al., 2011), with reduced white matter volume in adolescence (Kennedy et al., 2016), and with lower cerebral white matter integrity in older adults (Marks et al., 2011). Still other studies have examined large age spans ranging from very early adulthood into older age. These studies, too, have found that BMI negatively correlated with white matter integrity, especially in midbrain and brain stem tracts (Verstynen et al., 2012) and subregions of the corpus callosum (Stanek et al., 2011). Altogether, these findings have been used to argue that improving physical fitness and body composition might improve brain health, and in turn, promote cognitive performance during development (Chaddock-Heyman et al., 2015) and reduce cognitive impairment during aging (Erickson et al., 2014).

Analysis of links between physical fitness and body composition with neuroimaging markers of brain health specifically in early adulthood has been somewhat overlooked, despite acknowledgement that a lifespan perspective on brain health is warranted, in which risk factors for deterioration in brain health are potentially present across the lifespan (Moffitt et al., 2016;Williamson et al., 2018;Tucker-Drob, 2019). Evidence for such associations could provide the grounds for future research that explores whether intervening in young adulthood to promote fitness and body composition might alter long-term trajectories of brain health into older age.

The Young Adult Human Connectome Project (YA-HCP) is a large neuroimaging study of approximately 1200 women and men aged 22 to 36 (Van Essen et al., 2013), which offers an excellent data source for exploring links between physical fitness and body composition with various neuroimaging markers of brain health. Beyond collecting high-quality multi-model neuroimaging data, the YA-HCP collects behavioral data in a variety of domains, including physical fitness and body composition (Barch et al., 2013). Previous studies using the YA-HCP have observed that higher BMI and lower submaximal cardiovascular endurance are associated with lower integrity within several white matter tracts (Repple et al., 2018;Opel et al., 2019). Other YA-HCP analyses have shown that BMI correlates with cortical thickness, with the direction (negative versus positive) depending on the region and hemisphere (Vainik et al., 2018). In a recent study using the YA-HCP, we failed to observe an association between cortical thickness and either submaximal cardiovascular endurance or grip strength (Best, 2020). Like other studies in the field, our previous study was limited by focusing on only one type of brain structure (specifically, cortical thickness), despite knowledge that gray matter structure is distinct from white matter structure, and even that cortical thickness is genetically and phenotypically distinct from cortical area (Winkler et al., 2010). As such, it is plausible that fitness and body composition might have different associations with different structural features of the brain. Further, although our previous study adjusted for participant height and weight in the regression analyses, BMI was not considered as an important predictor in its own right, and it is possible for fitness and body composition to have unique associations with brain structure.

Thus, the current study is motivated in part by the need for a comprehensive analysis involving multiple aspects of physical fitness and body composition along with various structural measures of brain health within a single sample with sufficient sample size to appropriately correct for multiple testing while maintaining reasonable statistical power. Using data from the YA-HCP, the current study estimated associations between submaximal cardiovascular endurance, grip strength, and BMI with cortical gray matter thickness and area, with subcortical gray matter volume, and with white matter fractional anisotropy (FA), a commonly-used measure of white matter integrity based on the flow of water molecules along white matter tracts (Pierpaoli et al., 1996). Importantly, the default assumption made in most previous investigations is that associations between fitness/body composition and brain structure is linear across the range of scores; however, this may not necessarily be the case. Erickson and colleagues (2010) observed that the strongest association between physical activity and gray matter volume in older adults was at the highest quartile of physical activity, suggesting that a relatively large volume of physical activity might be necessary for detection of effects on brain structure. Whether such non-linear associations are observed for physical fitness or body composition is unknown, but the relatively large sample and wide distribution of fitness and body composition in the YA-HCP offers an opportunity to explore this possibility.

Another strength of the YA-HCP is the inclusion of genotype-confirmed monozygotic and dizygotic twin pairs, along with full-sibling non-twin pairs. Assuming that genetic overlap approximates 100% in monozygotic twins and 50% in dizygotic and non-twin pairs, behavioral genetic analyses can be conducted to estimate the heritability and environmental contributions to variation in fitness and brain structure, as well as the covariation between the two (Grasby et al., 2017). Previous research has found a moderate heritability contribution to the correlation between cardiovascular endurance and executive function (Best, 2020) and between BMI and executive function, cortical thickness and medial temporal lobe volume (Vainik et al., 2018). Research in other samples has shown a moderate heritability contribution to the BMI-executive function correlation in youth (Wood et al., 2019) and a more modest heritability contribution to the cardiovascular fitness-intelligence association among young men enlisted for military service (Aberg et al., 2009). Altogether, these studies suggest at least a modest genetic component to correlations between physical fitness and body composition, on the one hand, and neuro-cognition, on the other. To follow-up on our primary analyses, the current study conducted a set of behavioral genetic analyses to estimate the degree to which genetics or the environment underpin associations identified in the larger sample.

## Methods

### Study Design and Participants

Participant data were provided by the open-access YA-HCP 1200 Subjects Data Release (Van Essen et al., 2013). For our primary analyses with the entire sample, 1113 participants had valid Freesurfer-processed 3T structural MRI data and 1065 participants had valid FSL-processed diffusion imaging of white matter data. For behavioral genetic analyses, twin zygosity was verified by genotyping. Full-sibling, non-twin pairs were identified by having identifical mother and father identification numbers, but not being twins. Full-siblings with an age discrepancy ≥ 5 years were excluded from behavioral genetic analyses.^1^ For behavioral genetic analyses involving white matter FA values, data from 134 monozygotic twin pairs, 72 dizygotic twin pairs, and 290 full-sibling, non-twin pairs were available. For behavioral genetic analyses involving gray matter, data from 138 monozygotic twin pairs, 78 dizygotic twin pairs, and 313 full-sibling, non-twin pairs were available.

Participants were healthy adults within the age range of 22 to 36, who were drawn primarily from families that included twins in Missouri, USA. Families were excluded where individuals within the family had severe neurodevelopmental disorders (e.g., autism), neuropsychiatric disorders (e.g., schizophrenia) or neurological disorders (e.g., Parkinson’s disease). Individuals were also excluded if they had illnesses thought to impact neuroimaging data quality (e.g., high blood pressure or diabetes). Additional details on the participants and study design can be found elsewhere (Van Essen et al., 2013). Extensive details on all the measures included in the HCP study can be found at: https://wiki.humanconnectome.org. The original study was approved by the Washington University institutional review board. All participants provided written informed consent. Approval for this secondary analysis was provided by the institutional review board at Simon Fraser University (Title: “Genetic Contributions to Cognition and Physical Functioning in Young Adults: Analysis of the Human Connectome Project Study”; Study no: 2019s0471).

### Measures

#### Demographic and Other Covariate Variables

Age in years, gender, race/ethnicity, years of completed education, and annual total household income were self-reported. Gait speed (meters per second) was assessed over 4-meters, starting from rest, and was included to account for variance in the endurance measure due to differences in gait.

#### Physical Fitness

Participants completed several instruments from the NIH Toolbox for Assessment of Neurological and Behavioral Function on the first of the two-day assessment schedule (‘NIH toolbox’). The NIH toolbox includes psychometrically-validated measures of cognition and behavior, suitable for use across the human lifespan (Gershon et al., 2013; Reuben et al., 2013; Weintraub et al., 2013). Details on its implementation within the HCP can be found elsewhere (Barch et al., 2013).

Sub-maximal cardiovascular endurance was assessed by having participants walk as far as possible over a 2-minute period of time, back-and-forth over a 50-ft course. The total distance is recorded. Two trials were completed, with the faster trial used as the outcome of interest. Full force grip strength is measured with both hands using a Jamar Plus Digital dynamometer with the elbow bent to 90 degrees and arm against the trunk. Practice is conducted for each hand. Pounds of force for the dominant hand is used as the outcome of interest. Both fitness measures have shown good test-retest reliability (Intraclass correlation coefficient [ICC] ≥ 0.88) and convergent validity (r≥ 0.77) with ‘gold standard’ measures of endurance and grip strength across adulthood (Reuben et al., 2013). Further, although grip strength is a more direct measure of upper body strength, it also strongly correlates with knee extension performance (r≥ 0.77), a measure of lower body strength, suggesting grip strength may serve as a proxy body strength more generally (Bohannon, Magasi, Bubela, Wang, & Gershon, 2012). Height was reported in inches and weight was reported in pounds; these values were converted to body mass index with the following formula: 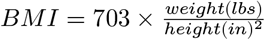.

### Structural MRI Data Acquisition and Processing

Gray matter imaging acquisition and processing was conducted by the YA-HCP research team with no additional processing on the freely-available data. Extensive details on the imaging protocol are found elsewhere, including details of additional scanning paradigms not included in the current study (Van Essen et al., 2012;Van Essen et al., 2013). All participants were scanned on a Siemens 3T customized ‘Connectome Skyra’ at Washington University in St. Louis, using a 32-channel Siemens head coil. Two T1-weighted and two T2-weighted scans were acquired at a spatial resolution of 0.7 mm isotropic voxels over two sessions. Each scan was evaluated by a trained rater for overall quality and only good/excellent scans were submitted to the structural pipelines, which used FreeSurfer 5.1 software and custom steps that combine the T1- and T2-weighted images for more accurate delineation of white and pial surfaces (Van Essen et al., 2013). Initial registration to MNI152 space was completed by FSL’s FLIRT tool, followed by nonlinear FNIRT to align subcortical structures. Cortical surface alignment was achieved using a surface-based registration using FreeSurfer, separately for each hemisphere. Average cortical thickness and surface area was obtained in 68 regions of interest (ROIs) across the two hemispheres. Additionally, subcortical gray matter volume was obtained in 14 ROIs in the limbic system.

Diffusion images were acquired at a spatial resolution of 1.25 mm isotropic voxels during a session including six runs (each 9 min 50 s in duration). Initial pre-processing of diffusion data was conducted by the YA-HCP research team and included intensity normalization across runs, ‘TOPUP’ susceptibility-induced distortion correction, and ‘EDDY’ eddy current and motion correction (Glasser et al., 2013). Subsequent processing was conducted by the current research team using standard tract-based spatial statistics in FSL (Smith et al., 2006). Fractional anisotropy (FA) images were registered to the FMRIB58 FA template and averaged to create a mean FA image. This image was used to create a WM skeleton using an FA threshold of 0.2. The Johns Hopkins University ICBM-DTI-81 white matter atlas (Mori et al., 2008;Oishi et al., 2008), which consists of 48 ROIs, was used to mask the WM skeleton and average FA values for every ROI was obtained per participant.

### Statistical Analyses

Analyses were conducted using R version 4.0.2 (r-project.org) using the Rstudio environment (Rstudio.com). An a priori power calculation was not conducted given that this is a secondary analysis of an existing dataset. For each analysis the maximum sample size possible was used. The first step was to regress the cortical thickness and area values, subcortical volumes and white matter FA values on a set of covariates (age, gender, ethnicity, income, education, gait speed, and intracranial volume) and the physical fitness measures (grip strength, submaximal endurance, and body mass index). All fitness measures were included together in the same regression model so as to model their independent associations with the neuroimaging outcomes. Generalized linear models fit by restricted estimated maximum likelihood were used for these regressions. To allow for nonlinearity in the association of age and physical fitness with the outcome of interest, restricted cubic splines were specified with knots located at the 10th, 50th, and 90th percentile values of the age and physical fitness variables (Harrell, 2019a). A compound symmetry covariance matrix accounted for the clustering of participants by maternal ID. A separate model was constructed for each outcome. All predictors were retained in the models regardless of the statistical significance of their Wald test. These regression models were constructed using `rms` version 6.0.1 (Harrell, 2019b). Control of the type I error rate was achieved by first allocating an equal portion of an overall α = 0.05 to each of the four outcome domains (i.e., cortical thickness and area, subcortical volume, and white matter FA) and then to each of the three key predictors (endurance, strength, and BMI). Next, because outcomes were anticipated to correlate with one another (i.e., thickness in one region correlates with thickness in another region), the effective number of independent tests within each outcome domain was calculated using an approach described by Nyholt (cite Nyholt 2004). Thus, the α for the test of a specific predictor and region within a domain was calculated as:

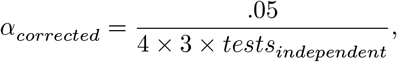

where the estimated number of independent tests was 40.6, 57.8, 52.4, and 10.3 for FA, cortical thickness, cortical area, and subcortical volume measures, respectively. These corrected two-sided α values translated into a Z statistic threshold of 3.88 for the FA outcomes, 3.97 for cortical thickness outcomes, 3.95 for cortical area outcomes, and of 3.54 for the subcortical volumes.

The second step was to estimate the proportion of phenotypic covariance between neuroimaging measures and physical fitness that can be attributed to heritability versus environmental sources. These analyses were limited to monozygotic twin, dizygotic twin, and full-sibling non-twin pairs with an age discrepancy of less than 5 years. Using the package `OpenMx` (version 2.17.4), a bivariate Cholesky decomposition twin model was constructed (Grasby et al., 2017). The Cholesky decomposition models specified that phenotypic variation in neuroimaging and physical fitness could be attributed to heritability variance (A), shared environmental variance (C), a twin-specific shared environment (T), and non-shared environmental variance (E) (**Figure 1**). These sources of variation are estimated by assuming that monozygotic twins share 100% of heritability effects and that dizygotic twins and full-sibling non-twins each share 50% of heritability effects. The shared environmental effects are equivalent for all three groups; additionally, monozygotic and dizygotic twins have another shared environmental effect that non-twins do not have. To estimate the heritability and environmental contributions to the correlation between neuroimaging and physical fitness, the observed physical fitness variable is also allowed to load onto the latent heritability and environmental (shared, twin-specific, and non-shared) contributors to neuroimaging variation (Grasby et al., 2017). The result is the estimation of the variation in neuroimaging and physical fitness, as well as the covariation between neuroimaging and physical fitness. Prior to being entered into the Cholesky decomposition, neuroimaging and physical fitness variables were regressed on age (with non-linear restricted cubic spline effects), sex, race, education, annual income, and the interaction between age and sex in an ordinary least squares regression model (one model per outcome including all twins together).

**Figure 1:**
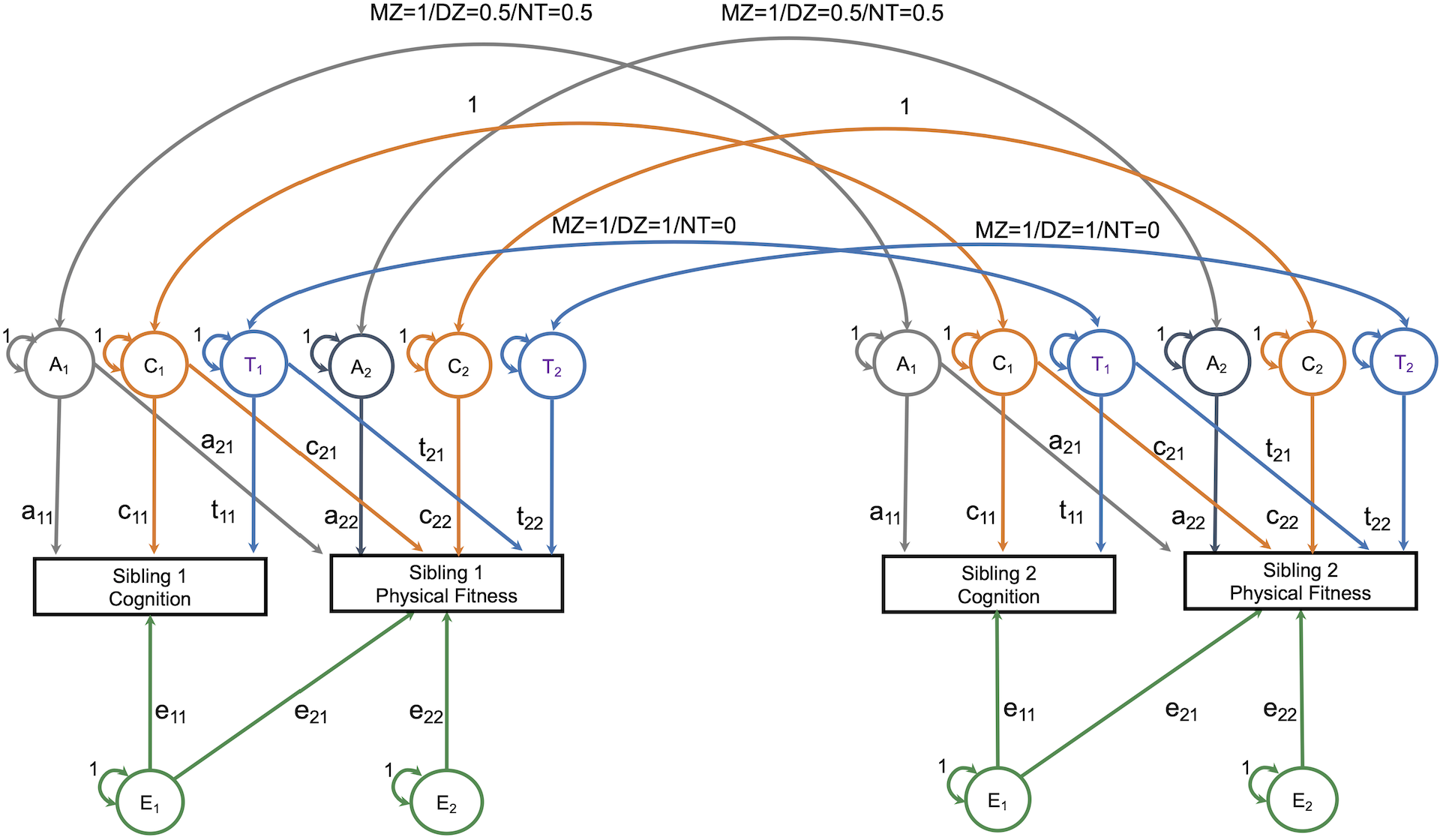
Overview of Bivariate Cholesky Decomposition Model

The assumptions of equality of means and variances across twin order and zygosity were tested using the likelihood ratio test of nested models. A saturated model was fit first, in which all means and variances were allowed to vary freely across twins and zygosity. Next, means and variances were constrained to be equal across twin order, and then, across zygosity. Finally, the ACTwE model was estimated. To summarize the results of this model, standardize estimates and standard errors for each of the labeled paths in **Figure 1** (e.g., “a11”) are presented, as well as the proportion of variation or of the cross-trait correlation that can be attributed to each of the components (i.e., A, C, Tw or E). (Note. Total variance or covariance is the sum of the A, C, Tw and E components. Covariance can then be converted into a correlation by dividing by the product of the standard deviation of the cognition measure and the standard deviation of the fitness measure).

All data used for the current study can be freely obtained by qualified registered researchers **here** and the supplemental material and complete R script can be found **here**.

## Results

### Sample Characteristics and Initial Associations

Demographic and physical fitness data on the full sample (i.e., all participants with GM and WM neuroimaging data) are summarized in *Table 1*. Mean age was roughly 29 years, and sample was 46% male. Nearly 70% of the sample was non-Hispanic white. Average body mass index fell in the overweight range. Endurance, and to a lesser extent grip strength, were on average higher than the age-based population norms (mean = 100) on which these measures were validated.

**Table 1.** Descriptive information on total sample

|  | Overall |
| --- | --- |
| n | 1065 |
| Age, yrs (mean (SD)) | 28.75 (3.67) |
| Sex, Male = M (%) | 490 (46.0) |
| Race/Ethnicity (%) |  |
| Asian or Pacific Islands | 64 ( 6.0) |
| Black/African American | 148 (13.9) |
| Hispanic/Latino | 94 ( 8.8) |
| Non-Hispanic, White | 734 (68.9) |
| Other | 25 ( 2.3) |
| Annual household income (%) |  |
| <\$10k | 72 ( 6.8) |
| 10k-20k | 82 ( 7.7) |
| 20k-30k | 136 (12.8) |
| 30k-40k | 126 (11.9) |
| 40k-50k | 106 (10.0) |
| 50k-75k | 225 (21.2) |
| 75k-100k | 146 (13.8) |
| >=100k | 166 (15.7) |
| Years of education (%) |  |
| 11 or less | 36 ( 3.4) |
| 12 | 146 (13.7) |
| 13 | 65 ( 6.1) |
| 14 | 131 (12.3) |
| 15 | 65 ( 6.1) |
| 16 | 454 (42.7) |
| 17 or more | 167 (15.7) |
| Gait speed (mean (SD)) | 1.32 (0.20) |
| BMI (mean (SD)) | 26.40 (5.11) |
| Endurance (mean (SD)) | 108.07 (13.94) |
| Grip strength (mean (SD)) | 103.56 (20.12) |

Simple bivariate correlations among age and physical fitness measures are shown in **Table 2**. Although gait speed was not a primary measure of interest, it is included as well. Age only weakly correlated with the other measures; endurance was modestly correlated with gait speed, grip strength and BMI (|*r*| ∼ 0.2 - 0.4).

**Table 2.** Correlations among physical fitness measures and age.

|  | Age | Endurance | Strength | BMI | Gait |
| --- | --- | --- | --- | --- | --- |
| Age | 1 | -0.09 | -0.09 | 0.09 | 0.04 |
| Endurance |  | 1 | 0.28 | -0.35 | 0.24 |
| Strength |  |  | 1 | 0.15 | -0.02 |
| BMI |  |  |  | 1 | -0.09 |
| Gait |  |  |  |  | 1 |

### Physical Fitness and Structural Neuroimaging Markers

#### White Matter Fractional Anisotropy

Twelve of the possible 48 white matter ROIs were associated with BMI with *z* test statistic that exceeded the multiplicity-corrected threshold. These ROIs are summarized in **Table 3** and are arranged from largest to smallest |*z*| statistic. In all cases, greater BMI was associated with lower FA values.

**Table 3.** Summary of effects of body mass index on white matter fractional anisotropy values

| White matter ROI | Abbr. | Effect | S.E. | Z stat |
| --- | --- | --- | --- | --- |
| Cerebral_peduncle_L | CpL | -0.0081 | 0.0011 | -7.7 |
| Fornix_cres_L | FcL | -0.01 | 0.0016 | -6.5 |
| Fornix_cres_R | FcR | -0.0092 | 0.0016 | -5.9 |
| Cerebral_peduncle_R | CpR | -0.0063 | 0.0011 | -5.8 |
| Pontine_crossing_tract | Pct | -0.0074 | 0.0015 | -5 |
| Sagittal_stratum_L | SsL | -0.0072 | 0.0015 | -4.9 |
| Cingulum_hippocampus_R | ChR | -0.0094 | 0.002 | -4.7 |
| Retrolecticular_part_internal_capsule_R | RpicR | -0.0062 | 0.0014 | -4.4 |
| Cingulum_hippocampus_L | ChL | -0.0082 | 0.0019 | -4.3 |
| Corticospinal_tract_L | CtL | -0.0073 | 0.0018 | -4.1 |
| Superior_frontooccipital_fasciculus_R | SffR | -0.0057 | 0.0015 | -3.9 |
| Inferior_cerebellar_peduncle_R | IcpR | -0.0072 | 0.0018 | -3.9 |

Marginal plots of the association between BMI and these white matter ROIs are shown in **Figure 2** below, which account for the covariates and other fitness measures. In some instances, the association between BMI and FA was fairly constant across the range of BMI values (e.g., left and right fornix), whereas in other instances, the association appeared to dissipate somewhat with more extreme BMI values (e.g., left and right cerebral penduncle).

**Figure 2:**
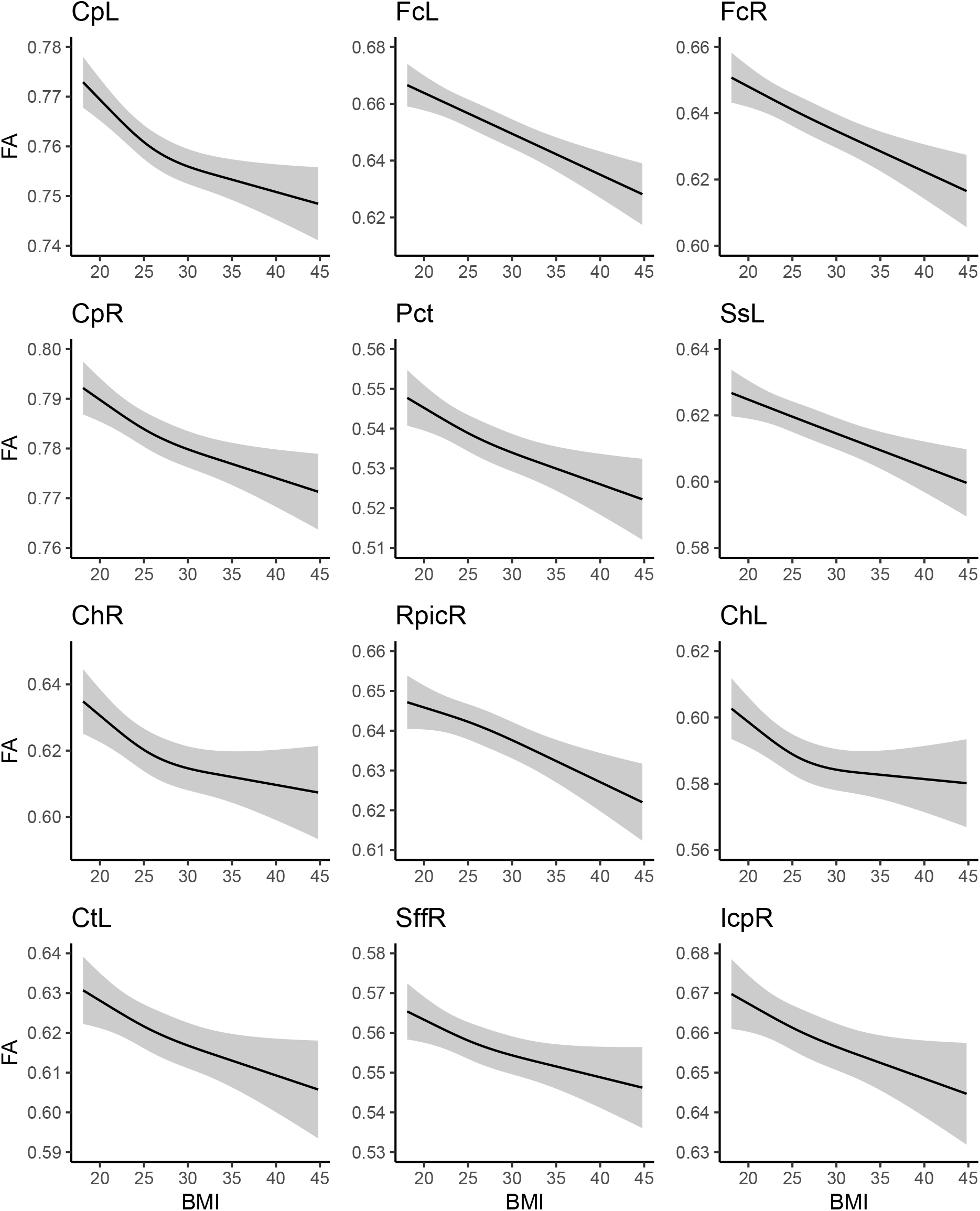
Association between BMI and white matter FA values

Neither endurance nor grip strength had associations with the white matter ROIs with *z* statistics beyond the threshold. Comprehensive results for all white matter ROIs can be found in section 1 for the supplemental material.

#### Gray Matter Structure

Body mass index was positively associated with right superior parietal thickness (see **Table 4**). As shown in the marginal plot in **Figure 3** this association was strongest as BMI increased from the normal to the overweight range and then weakened somewhat with more extreme BMI values. No additional fitness associations with gray matter structure (whether thickness, area, or subcortical volume) had significant *z* statistics. Comprehensive results for all gray matter ROIs can be found in section 2 for the supplemental material.

**Table 4.**
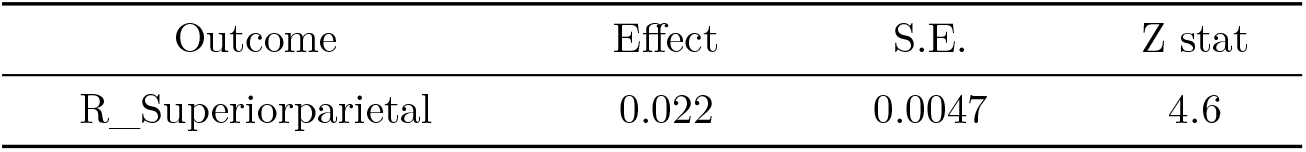
Summary of effects of body mass index on GM cortical thickness

| Outcome | Effect | S.E. | Z stat |
| --- | --- | --- | --- |
| R_Superiorparietal | 0.022 | 0.0047 | 4.6 |

**Figure 3:**
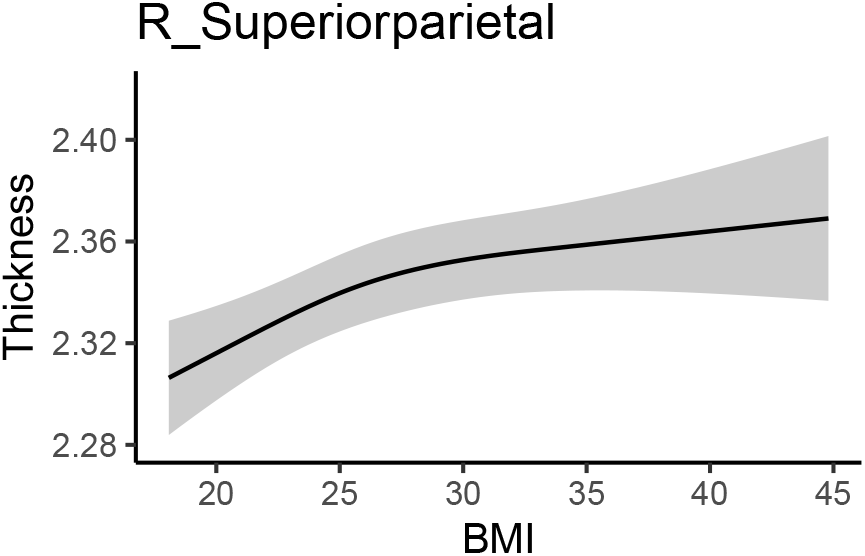
Association between BMI and GM thickness values

### Bivariate Behavioral Genetic Analysis of Fitness and Brain Structure

Based on the results of the larger sample, bivariate genetic models were estimated between body mass index and 12 white matter ROIs and right superior parietal gray matter thickness. The full ACTwE model showed small estimates with large standard errors for the shared environmental and twin-specific environmental effects. As has been done in previous research (Wood et al. 2019;Vainik et al. 2018), the loadings and cross-loadings for these two effect were fixed to zero, resulting in a simpler AE bivariate model. Across brain regions, this simpler AE model did not result in a worsening in fit according to likelihood ratio tests, and therefore, is described below. Comparison between ACTwE and AE models and tests for the assumptions of equality of means and variances can be found in section 3 of the supplemental material.

#### White Matter Fractional Anisotropy

Initial bivariate correlations across siblings, across traits, and across siblings and traits for the the three sibling pair types are shown in **Figure 4** for the 12 white matter ROIs. These provide a helpful preview to the formal behavioral genetic analyses presented below. Body mass index was strongly correlated among MZ twins (*r* ∼ 0.61), and was weakly correlated among DZ twins and non-twin siblings (*r* ∼ 0.20 - 0.27). This suggests an important role of heritability in BMI variation. Similarly, white matter FA was most strongly correlated within MZ twins for all 12 ROIs, also suggesting an important role of heritability for white matter integrity. Whether DZ twins or non-twin siblings had the next strongest correlation varied by ROI, thereby suggesting inconsistency with regard to twin-specific versus a more general shared environmental effect. Cross-trait and cross-trait/cross-sib (‘ct/cs’ in **Figure 4**) correlations also varied somewhat depending on the white matter ROI, though in general, the strongest negative correlation was among MZ twins and further, the cross-trait/cross-sibling correlation was nearly as strong as the cross-trait correlation among MZ twins. This implies at least some role of heritability in the correlation between BMI and FA values; however, in light that these correlations are generally quite modest (i.e., at most *r* ∼ −0.30), the absolute role of heritability is anticipated to be modest.

**Figure 4:**
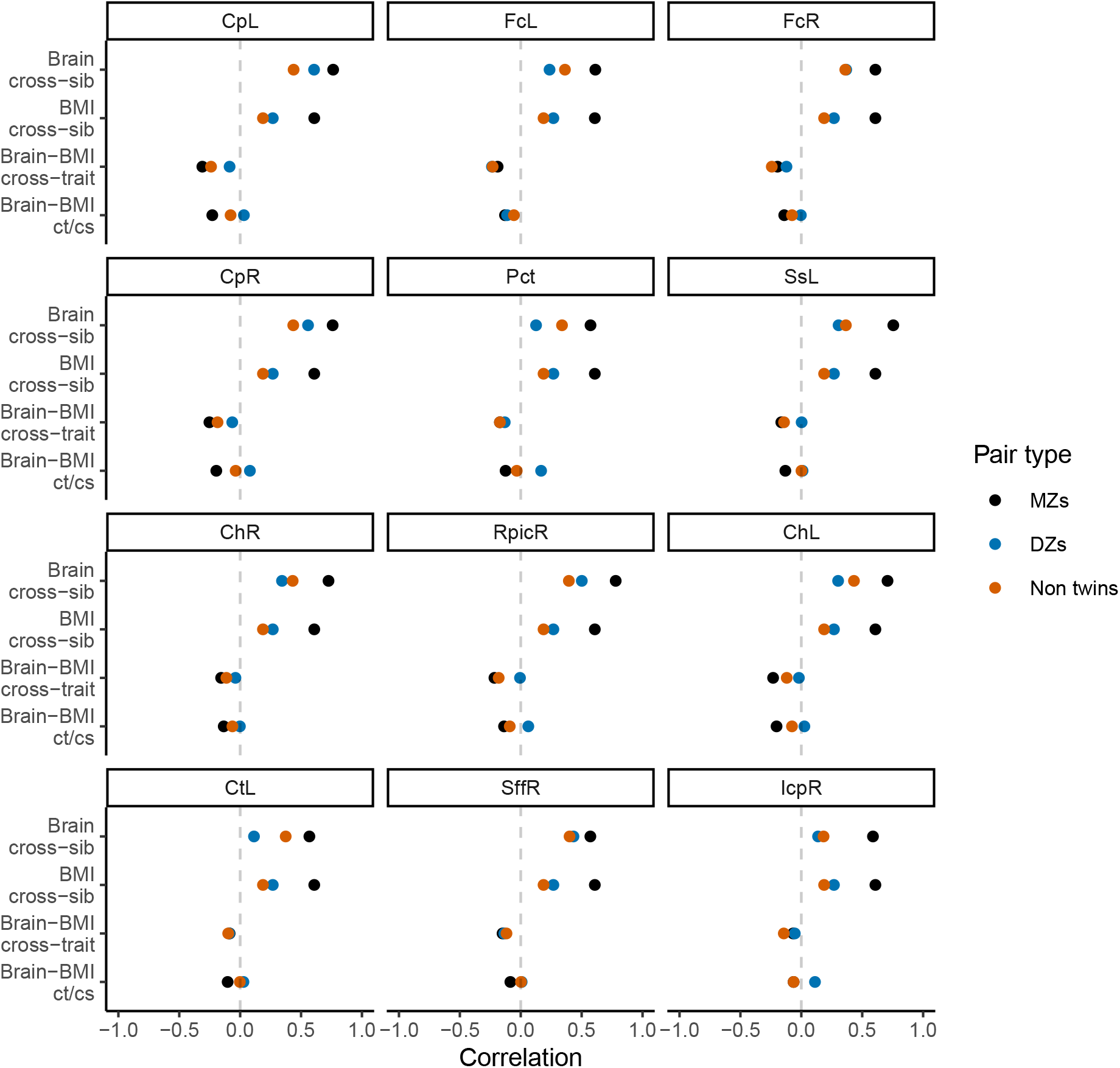
Phenotypic correlations across siblings and across traits

Standardized estimates and 95% confidence intervals for each of the paths in the bivariate cholesky decomposition model are provided for each of the 12 white matter ROIs in **Figure 5**. Following on the correlations described directly above, there were strong heritability loadings on white matter FA variance (path a11) and BMI variance (a22). Similarly, there were strong, precisely estimated environmental effects for FA variance (e11) and BMI variance (e22). The point estimate for the genetic cross-loading (a21) was negative and modest (*β* ∼ −0.20); in some instances, the confidence interval for this estimate crossed into positive numbers, suggested some uncertainty about the nature of this effect. The point estimate for the environmental cross-loading (e21) was also quite small, generally smaller in magnitude than the genetic cross-loading. The confidence interval for this estimate was fairly narrow, but at times crossed into positive numbers.

**Figure 5:**
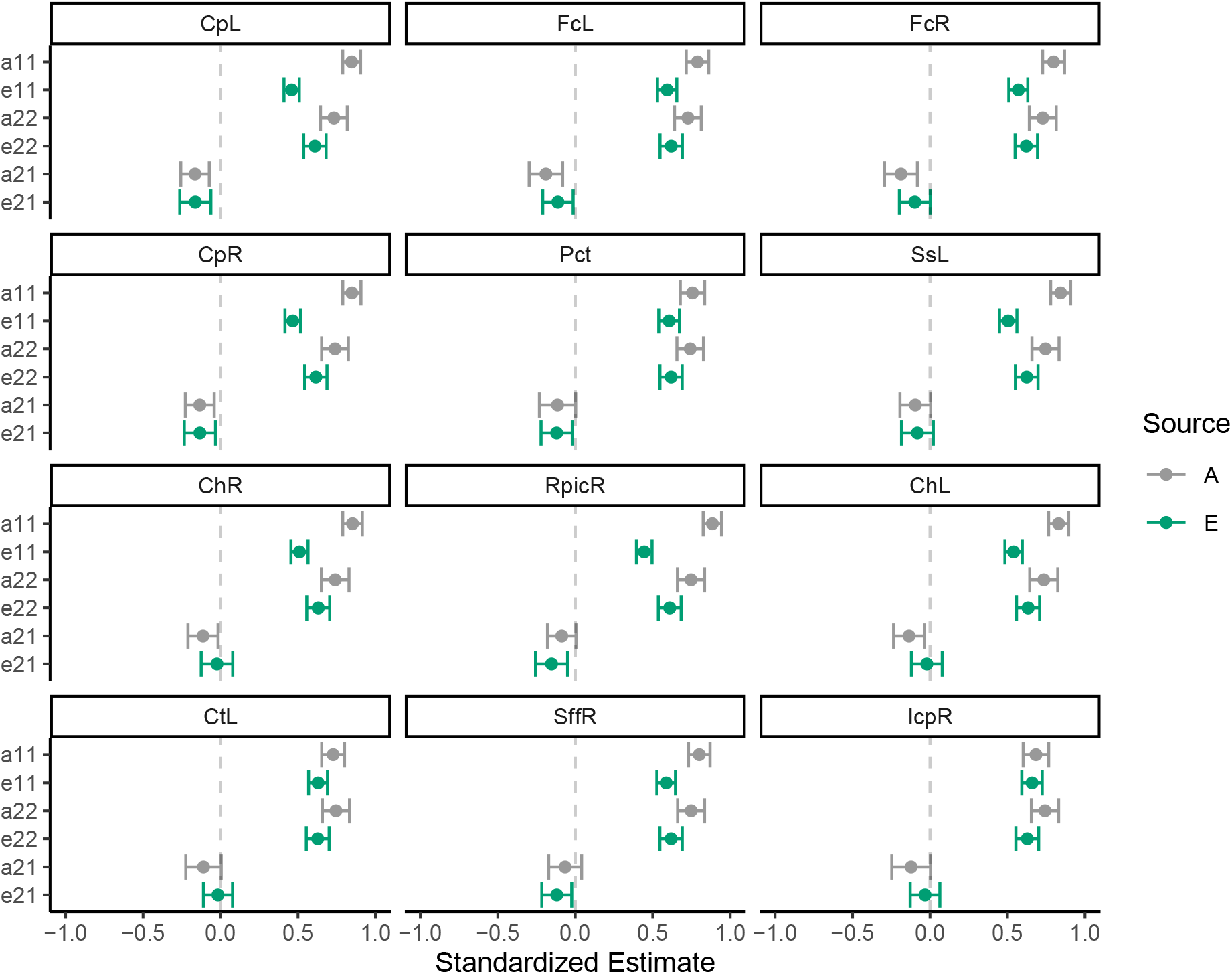
Behavioral genetic path estimates for BMI and white matter FA

These standardized estimates were then converted into variances and used to visualize how much of variation in FA values and BMI could be attributed to each of the four sources. For example, in the AE model, white matter FA variance is defined as below:

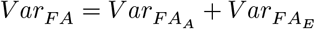

For white matter FA, each of the variance components is the square of the respective standardized loading For example,

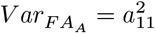

For BMI, each of the variance components is the sum of the respective squared cross-loading and the squared endurance-specific standardized loading. For example,

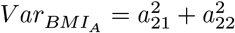

The estimates from the AE model can also be used to partition the phenotypic correlation into A and E components. For example, the portion of the correlation attributed to heritability can be estimated by first estimating the covariance:

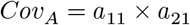

And then coverting that value into a correlation.

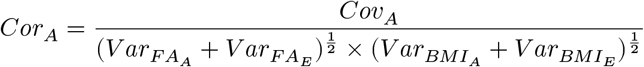

Proportions of the variances and of correlations are plotted in **Figure 6**. The decomposition of BMI variance is roughly identical across models (as expected) and shows that heritability contributed to roughly 50% of variance. Similarly, heritability contributed to between 50%-78% variance in white matter FA, depending on the ROI. Environmental factors, by design, explained the remainder of the variance. Covariances were converted to correlations and are also shown in **Figure 6**. As expected the total phenotypic correlation was modestly negative, ranging between −0.20 and −0.25. Generally, heritability accounted for 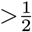 of this correlation. Section 4 of the supplemental material shows the analysis of these 12 white matter ROIs using the ACTwE model.

**Figure 6:**
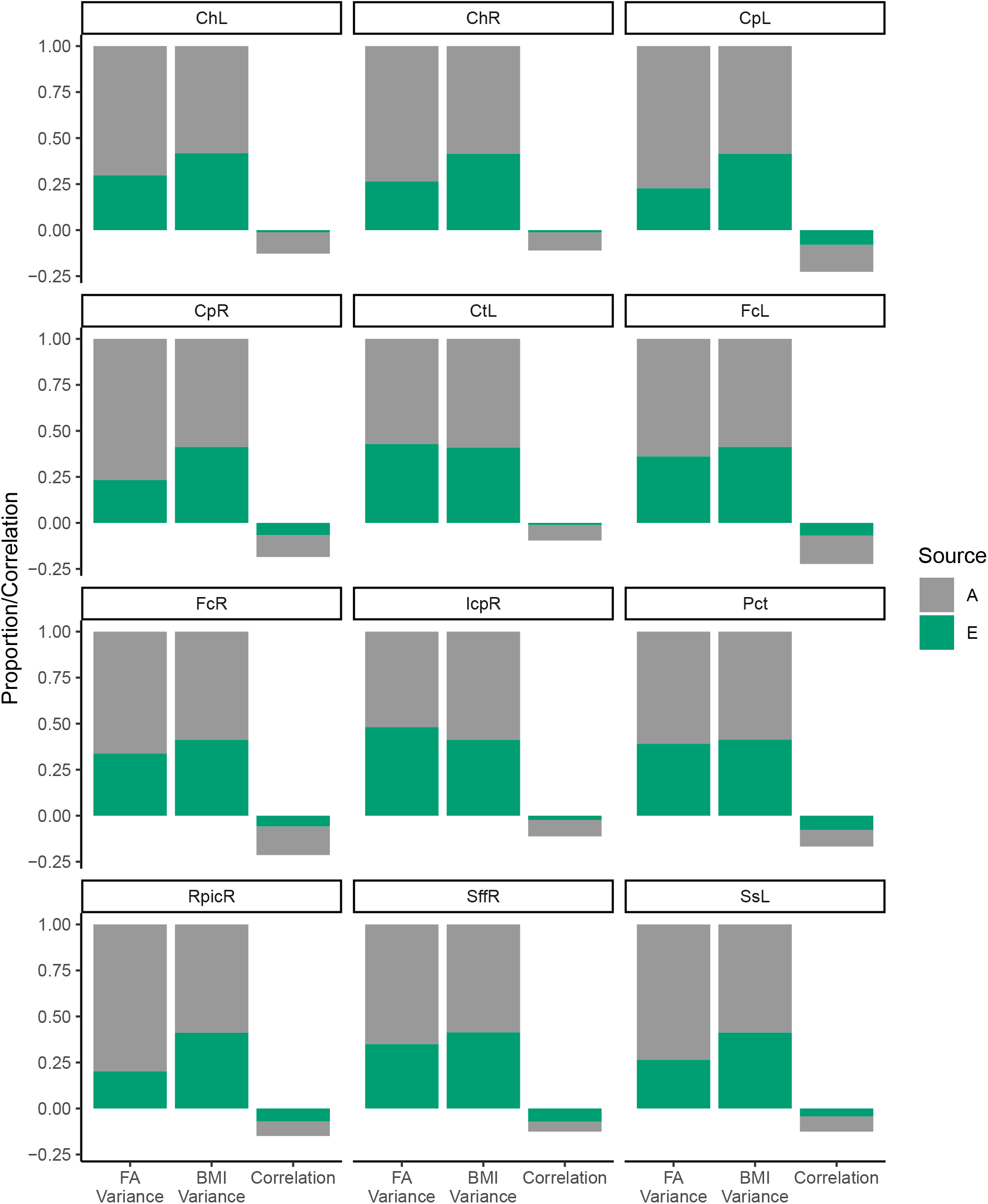
Variance components for BMI and white matter FA

#### Gray Matter Structure

All of the above behavioral genetic analyses were repeated on the right superior pariatal thickness, with the results summarized in **Figure 7**. As can be seen across the panels, there was a strong heritability effect for cortical thickness in addition to BMI (as described above). Analysis of the contributors to the correlation between BMI and cortical thickness in this region showed small positive effects for heritability (a21) and environment (e21) with confidence intervals that slightly cross into negative territory (see panel B). Heritability was estimated to contribute to 74% of variation of the variance in gray matter thickness and to slighly over 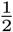of the small positive phenotypic correlation (panel C). Section 5 of the supplemental material shows the analysis of this gray matter ROI using the ACTwE model.

**Figure 7:**
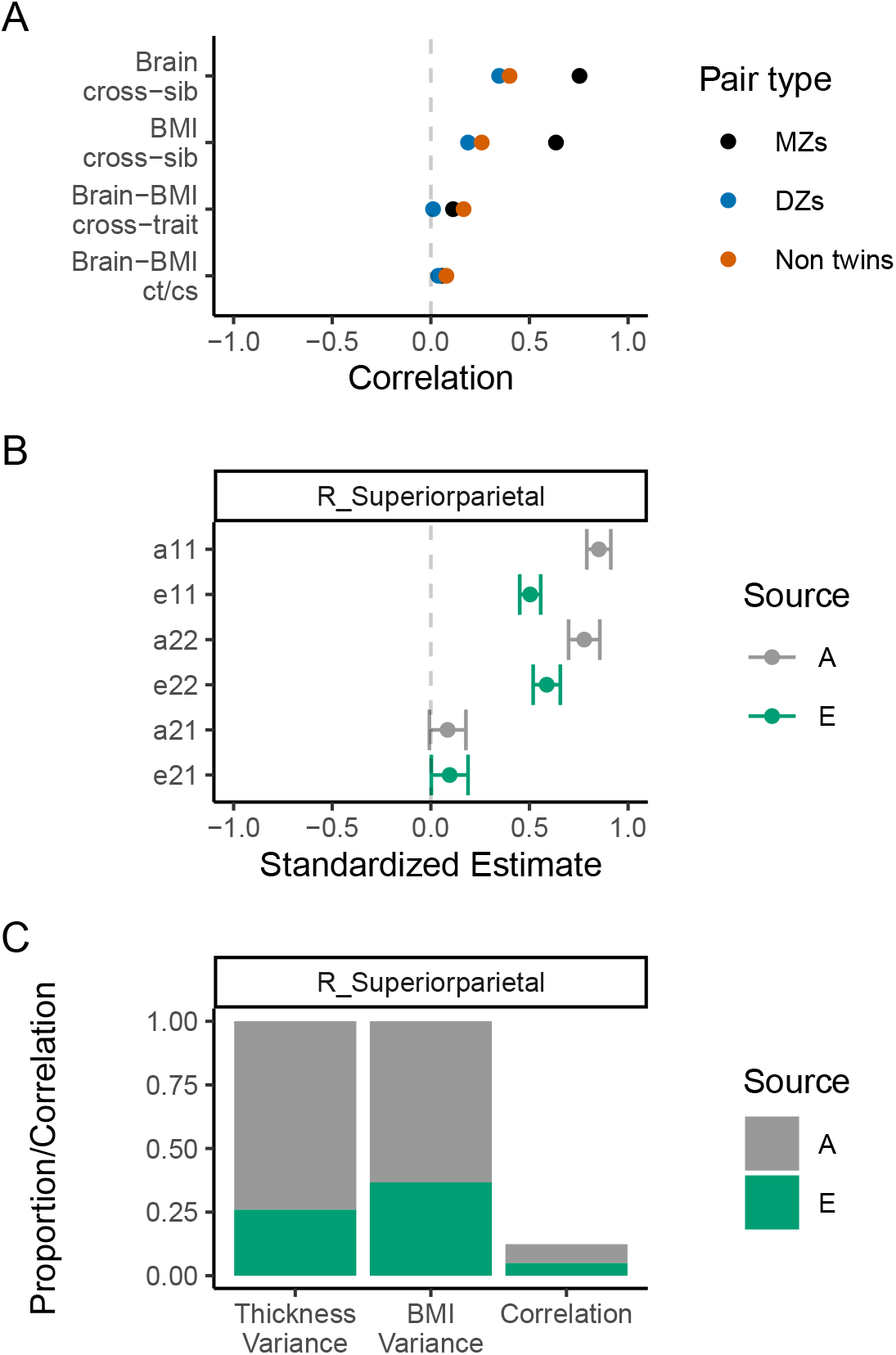
Summary of behavioral genetic analyses of BMI and right superior parietal thickness

## Discussion

In this large study of young adults, we observed that BMI, but not performance-based measures of fitness, negatively correlated with white matter integrity in various white matter tracts. Except for a single positive association between BMI and cortical thickness in the right superior parietal lobe, we failed to observe associations between either physical fitness or BMI with cortical or subcortical gray matter structure.

### Body Composition and Neurocognitive Health

These results add to a growing literature associating high BMI and obesity with lower neurocognitive functioning and brain health at various points in the lifespan (Gunstad et al., 2007;Gunstad et al., 2008;Raji et al., 2010;Kennedy et al., 2016). As expected, our results confirm a previous study that included data from the YA-HCP that showed that high BMI was associated with widespread reduced white matter integrity (Repple et al., 2018). The novelty of the current findings is that by also including several structural markers of gray matter, we observed a fairly selective association between BMI and white matter integrity that does not extend to gray matter, with the single exception in the right superior parietal lobe. Also, by including upper-body strength and submaximal cardiovascular endurance as additional predictors of interest, we show that the effects of BMI cannot be explained by these performance-based fitness measures, and further, that these fitness measures were not uniquely associated with brain structure. Finally, by allowing for nonlinearities in these associations, we observed that in instances in which the association deviated from linear, the most common pattern was for the slope to be steepest over the range of BMI values representing normal through overweight (i.e., 18 to 30), after which the slope became shallower. This was most apparent for white matter in the bilateral cerebral peduncle and bilateral hippocampal portion of the cingulum, and for the right superior parietal gray matter thickness.

Our findings are consistent with previous studies showing that although there is a negative association between BMI and white matter integrity throughout the brain, among the strongest statistical signals were subcortical brain stem pathways, including bilateral cerebral peduncle and pontine cross tract (Verstynen et al., 2012;Repple et al., 2018). By linking the midbrain to the thalamus, and to the cerebrum more generally, the cerebral peduncles are critically involved in motor and sensory functions, including motor control and coordination (Jane et al., 1968). Our findings also support previous studies showing a negative association between BMI and white matter integrity in the fornix (Stanek et al., 2011;Xu et al., 2013). The fornix carries projection fibers from the hippocampus and is implicated in higher-order learning and memory (Kantarci, 2014). Decreased fornix FA is observed with increasing age, and in mild cognitive impairment and Alzheimer’s disease (Kantarci, 2014). We also observed the BMI-FA association in the bilateral hippocampal portion of the cingulum, which has also been linked to memory functioning (Ezzati et al., 2016) and to Alzheimer’s disease (Dalboni da Rocha et al., 2020). In light of the association of BMI in midlife with risk of dementia in late life (Kivimaki et al., 2018), it is possible that these white matter tracts may play some role in transmitting this effect.

Similar to a previous analysis of YA-HCP data (Vainik et al., 2018), we observed a positive association between BMI and right superior parietal thickness; however, unlike this previous analysis, we failed to observe associations within other gray matter regions. Reasons for this discrepancy may include a stricter significance threshold in the current study that did not detect the weaker effects reported previously and the larger set of covariates used in the current study, which notably included performance-based physical fitness, educational attainment, and annual income.

We also failed to observe associations between either submaximal cardiovascular endurance or grip strength with any of the markers of brain structure, which is inconsistent with a previous analysis of the YA-HCP data that found an association between endurance and white matter integrity (Opel et al., 2019). However, the current analysis differed in its analytic approach by including a larger set of covariates and using an ROI-based, as opposed to a voxel-based, approach to analyzing the associations between BMI and neuroimaging data. The lack of associations between fitness and brain structure is also inconsistent with other studies that have explored these associations in youth (Chaddock-Heyman et al., 2015) and older adults (Oberlin et al., 2015). The inconsistency with the current results could arise for numerous reasons. It is possible that these associations are weaker during young adulthood as compared to during childhood development or aging. Previous studies (Chaddock-Heyman et al., 2015;Oberlin et al., 2015) also used superior measures of fitness (e.g., direct assessment of maximal oxygen uptake during a graded exercise test), which may have provided better measurement of fitness, and in turn, of the true association with brain structure. It is also possible that the large sample size of the current study allowed us to avoid detecting false positive findings. An underappreciated phenomena is that small studies, in which constructs are measured with error (as is ubiquitous in the behavioral and neurocognitive sciences), are prone to overestimate effect sizes, leading to false positives, in addition to being prone to failure in detecting effects (Loken and Gelman, 2017). This is especially true when the publication of research findings is contingent of the findings being statistically significant. To resolve the inconsistencies of the current study with previous ones, large-sample, lifespan studies with gold-standard measures of physical fitness are needed; the result will be better estimation of the associations between fitness and brain structure and determination of how these associations might change over the life course.

### Behavioral Genetics of Body Composition and Brain Structure

To further explore the nature of the association between BMI and brain structure, our study made use of the large set of twins and full siblings contained in the YA-HCP. Using behavioral genetic analyses, we estimated the degree to which variation in BMI and brain structure, and the covariation between BMI and brain structure, could be explained by heritability—i.e., a set of common genes that directly or indirectly impact BMI and brain structure—versus environmental effects. The first insight from these analyses was that a simpler behavioral genetic model, in which variation and covariation was partitioned into genetic versus environmental effects, fit the data no worse, despite being simpler, than a model which further apportioned environmental effects into twin-specific effects, more generally sibling shared effects, and non-shared environmental effects. Others have also found that this simpler model fits (co)variation in BMI and neurocognition well (Vainik et al., 2018;Wood et al., 2019). This would imply that shared environmental effects—whether specific to twins or to full siblings more generally—have little consistent contribution to any connections between BMI and brain structure.

The second insight from these behavioral genetic analyses is that there was a clear heritability effect underpinning variation in BMI and the identified brain structures, as well as the covariation between BMI and these brain structures. Indeed, this heritability effect exceeded the environmental effect across all variance and correlation estimates (see Figure 6 and Figure 7c) and comports with previous studies that have observed a clear genetic effect on variation in BMI and neurocognition analyzed independently (Friedman et al., 2008;Elks et al., 2012;Kochunov et al., 2015), as well as in studies that employed bivariate behavioral genetic analyses similar to the current study (Vainik et al., 2018;Wood et al., 2019).

This heritability effect is the portion of the variance or covariance that can be attributed to an additive genetic effect after adjusting for the covariates. Although heritability estimates provide some insight as to how various phenotypes (say, BMI and brain structure) are related, there is still a large degree of ambiguity about the nature of these associations. This clear heritability effect could reflect a common set of genes that causally affect both BMI and brain structure, so-called genetic confounding or pleiotropy (Vainik et al., 2018). If genetic confounding largely accounts for the association between BMI and brain structure, intervening upon either phenotype would be expected to have little impact on the other phenotype. Alternatively, genetic effects could directly affect one phenotype that in turn, through active selection of environments and behaviors, impacts the second phenotype (Scarr and McCartney, 1983). If our heritability estimate truly reflects a cascading effect where genetic effects are transmitted through an intermediary phenotype, there is evidence in the literature that the cascade could flow in either direction. One possibility is that differences in brain structure impact obesogenic behaviors through decision-making processes, and in turn, leads to variation in BMI (Alonso-Alonso and Pascual-Leone, 2007;Lowe et al., 2019). Under this proposed directionality, it may be helpful to remediate decision-making processes in order to promote healthful behavior or structure the environment to support healthful behaviors and reduce the reliance of individual decision-making (Hall and Fong, 2015;Hall, 2016), all with the intent of reducing obesity. Alternatively, BMI might impact brain structure via cardiometabolic effects, including inflammation and hypertension (Williamson et al., 2018;Repple et al., 2019). Interventions to reduce BMI might have the added benefit of improving brain structure, especially white matter integrity, though evidence from clinical trials is lacking (Wassenaar et al., 2019). It may also be worthwhile to intervene on the cardiometabolic consequences of excess weight. A recent clinical trial showed that intensive blood pressure treatment reduced the accrual of white matter lesions over time among hypertensive adults 50 years or older (Nasrallah et al., 2019)

## Limitations

The current study has noteworthy limitations and the findings are conditioned on certain untested assumptions. The cross-sectional study design prohibits us from understanding the causality or even directionality of the association between BMI and brain structure. As noted above, even a strong heritability effect is consistent with various causal models of the studied variables. Our physical fitness and body composition measures also have limitations. Physical fitness was measured less precisely than gold-standard measures, such as maximal oxygen uptake during a graded exercise test. BMI was calculated using self-reported height and weight. Our behavioral genetic analyses require the untested assumptions that there is no assortative mating (i.e., individuals are not mating with others with similar BMI or brain structure more than expected at random) and that the environmental effects are equivalent across sibling pair type.

## Conclusions

White matter integrity begins to decline in mid adulthood and it correlates with cognitive performance; furthermore, loss of white matter integrity and other white matter abnormalities are features of various dementia subtypes, including Alzheimer’s disease (Wassenaar et al., 2019). Our study adds to a fairly consistent pattern of findings showing that higher BMI correlates with lower white matter integrity at various points in the lifespan (summarized in Wassenaar et al., 2019). Most of these previous studies have examined mid or later adulthood. Along with a small set of previous studies (Aberg et al., 2009;Repple et al., 2018;Williamson et al., 2018;Opel et al., 2019;Repple et al., 2019), we bring attention to the fact that modifiable risk factors, such as BMI, correlate with brain health even in young adulthood. Our results support a lifespan perspective on neurocognitive aging, in which it may be fruitful to intervene many decades prior to the onset of cognitive impairment to effectively reduce negative impacts of cognitive aging (Moffitt et al., 2016;Tucker-Drob, 2019).

We also extend the current literature in other ways. By taking a comprehensive approach to analyzing fitness and body composition with several structural markers of brain health, we show that BMI, but not strength or endurance, uniquely correlates with white matter integrity, and that there was very limited evidence for an association with gray matter structure. A final contribution of the current work stems from findings from our behavioral genetic analyses of twins and full siblings. These analyses confirm previous studies by showing that heritability contributes to a majority proportion of variation in BMI and brain structure. Furthermore, we show that generally at least 1/2 of the correlation between BMI and brain structure can be attributed to heritability. This does not imply that either BMI or brain structure is immutable, but it does improve our understanding of the etiology of the association between BMI and brain structure.

## Data Availability

All data used for the current study can be freely obtained by qualified registered researchers (https://db.humanconnectome.org/) and the complete R analysis script and supplemental results can be found at https://osf.io/ykcm6/.

https://osf.io/ykcm6/

## Author Note

Data were provided by the Human Connectome Project, WU-Minn Consortium (Principal Investigators: David Van Essen and Kamil Ugurbil; 1U54MH091657) funded by the 16 NIH Institutes and Centers that support the NIH Blueprint for Neuroscience Research; and by the McDonnell Center for Systems Neuroscience at Washington University.

## Footnotes

1 It was possible for a given individual to be included in multiple sibling pairs, as there were families with up to six full siblings. In such instances, values were averaged across all pairs of the same type (i.e., DZ, MZ, non-twin full-sibling types) in which the given individual was involved, so as to avoid double-counting individuals. The vast majority of these involved non-twin, full-sibling pairs.

